# The Water, Sanitation, and Hygiene (WASH) for Everyone controlled before-and-after (CBA) trial: trial protocol and baseline results

**DOI:** 10.1101/2024.10.16.24315614

**Authors:** Kondwani Chidziwisano, Mindy Panulo, Clara MacLeod, Marcella Vignieri, Blessings White, Ian Ross, Tracy Morse, Robert Dreibelbis

## Abstract

Community-based behaviour change interventions are a common approach to Water, Sanitation, and Hygiene (WASH). Yet, published evaluations of how these interventions work in district-wide approaches are rare. This study reports the baseline characteristics and study design for a trial assessing the effectiveness of a district-level Community-led Total Sanitation (CLTS) intervention compared to the additional integration of local Care Groups on sanitation coverage and use and hygiene behaviours in Chiradzulu District, Malawi.

This study is a controlled before-and-after trial with two treatment arms and a control group. Clusters are rural villages in three Traditional Authorities (TAs). One arm receives CLTS and the Care Group Model, one arm receives CLTS only, and one serves as the control group. The trial is part of the wider WASH for Everyone (W4E) project, led by World Vision Malawi which aims to expand access to WASH services across the entire district by 2025. Study participants were selected from the three TAs. Systematic sampling procedures were used to select 20 households per cluster with a total of 1,400 households at both baseline and endline. The primary outcome is sanitation coverage. Secondary outcome measures include sanitation use, safe disposal of child faeces, observed handwashing facility, and Sanitation-related Quality of Life index (SanQoL-5).

Baseline results show a no difference for primary and secondary outcomes between arms. We noted low coverage of handwashing facilities with soap and water in all the three arms (i.e., CLTS only = 7%; CLTS and Care Groups = 4%; and control = 10%). Further, there was a slight variation (p= 0.08) in handwashing practice and sanitation coverage among the study arms.

The baseline observations indicate a balanced distribution of potential demographic confounders in the trial arms. The trial intervention is under implementation. The evaluation report is expected to be published in 2025.

## Introduction

Globally, it is estimated that 3.6 billion people lack access to basic sanitation services and 494 million people practice open defecation (OD), with the highest rates of OD in sub-Saharan Africa. (1). Consequences of OD include faecal contamination of drinking water sources and food, which contribute to a high burden of diarrhoeal diseases and child stunting, adversely impacting health and socio-economic development (2,3). Further, OD and inadequate sanitation disproportionately affect the safety and dignity of women, girls and marginalised groups (3–6), as well as other aspects of quality of life (7). Efforts by governments and other sanitation stakeholders to eliminate OD, such as the provision of subsidised latrines to households combined with hygiene and health education programmes, have failed to make adequate sustained progress (8,9). Behaviour centred interventions have been associated with improved uptake of sanitation interventions, but more information is needed to assess their impact on behavioural outcomes when implemented in combination with one another.

Our study focuses on two specific community-led interventions widely used in the WASH sector. The first is Community-Led Total Sanitation (CLTS), an approach to sanitation behaviour change centred on community-wide behaviour change and community self-enforcement in rural settings (Kar & Chambers, 2008). Introduced in Bangladesh in 2009 and now adopted globally, the major goal of CLTS is to mobilize communities to construct and use latrines to end OD (10). CLTS uses three phases to leverage social and emotional drivers to “trigger” a change in people’s mindsets towards OD (11). Evidence on the efficacy of CLTS is mixed. Certain studies highlight that CLTS only generates significant short-term impact for reducing OD through increased latrine coverage and use (9,12–15). CLTS implementation factors, such as triggering session attendance, number of supportive community leaders, participant’s anticipation to receive an incentive, and the number of follow-up visits, have been reported to significantly influence latrine coverage (16). The second is the Care Group (CG) model which relies on a multiplier effect to reach a high number of households in a community at low-cost through a development of a supportive network of peer-to-peer counselling (17).The CG model is a well-tested programme for the delivery of health interventions in rural communities, historically focusing on maternal and child health (17–20). Twenty-three non-governmental organisations (NGOs) have implemented the CG Model across 27 countries, including Malawi (18,20). Studies have documented the effectiveness of the CG Model in increasing coverage of child survival interventions and reducing under five mortality (19–21). However, despite their clear alignment, studies of the effectiveness of CGs as they relate to WASH interventions are limited.

In Malawi, communities struggle to sustain 100% latrine coverage after attainment of Open Defaecation Free (ODF) status (22). This has been attributed to a number of factors, including lack of involvement of marginalized and disadvantaged people, use of low-quality building materials, lack of technical support and improper programme implementation (12,23). To achieve high and sustained latrine coverage and behaviour change, it is essential to address all physical and contextual factors that directly relate to long-term CLTS success. The Government of Malawi adopted CLTS as one of its official approaches for sanitation in 2008 (24). The Government formally adopted the CG model in 2011 as an operational framework for the Scaling Up Nutrition (SUN) Strategy (25). However, little is known about how effective the CG model can be in promoting wider community health benefits, such as improved sanitation. Models to promote sustained reductions in open defaecation and improved sanitation outcomes through combination of behaviour centred intervention need to be tested and adopted to support long-term positive health outcomes.

This study aims to assess the effectiveness of Community-led Total Sanitation (CLTS) combined with the Care Group model on sanitation coverage and use and hygiene behaviours in Chiradzulu, District, in rural Malawi, compared to CLTS alone or no intervention. The study objectives are to assess: 1) how the two interventions compare to one another for improving sanitation coverage and use, and 2) whether the two interventions are individually more effective than no intervention at all.

## Methods and analysis

### Study setting and population

The study is implemented in Chiradzulu District, Malawi (Fig 1). Chiradzulu District is situated in the southern region of Malawi and is sub-divided into 10 administrative regions, or Traditional Authorities (TA). The Malawi 2015/16 Demographic and Health Survey (DHS) indicated that 52% of the population has access to improved sanitation. According to the National Statistical Office (NSO), in 2019, 93% and10% of the households in Chiradzulu District had access to safe water and improved sanitation facilities, respectively. OD rates in Malawi and Chiradzulu District are 6% and 7%, respectively (26). Given this low level of coverage, Chiradzulu District is the target of a three-year (2021–2024) district-wide water, sanitation and hygiene programme, known as WASH for Everyone (W4E), implemented by World Vision and Water for People, alongside which this trial is embedded.

**Figure 1.**
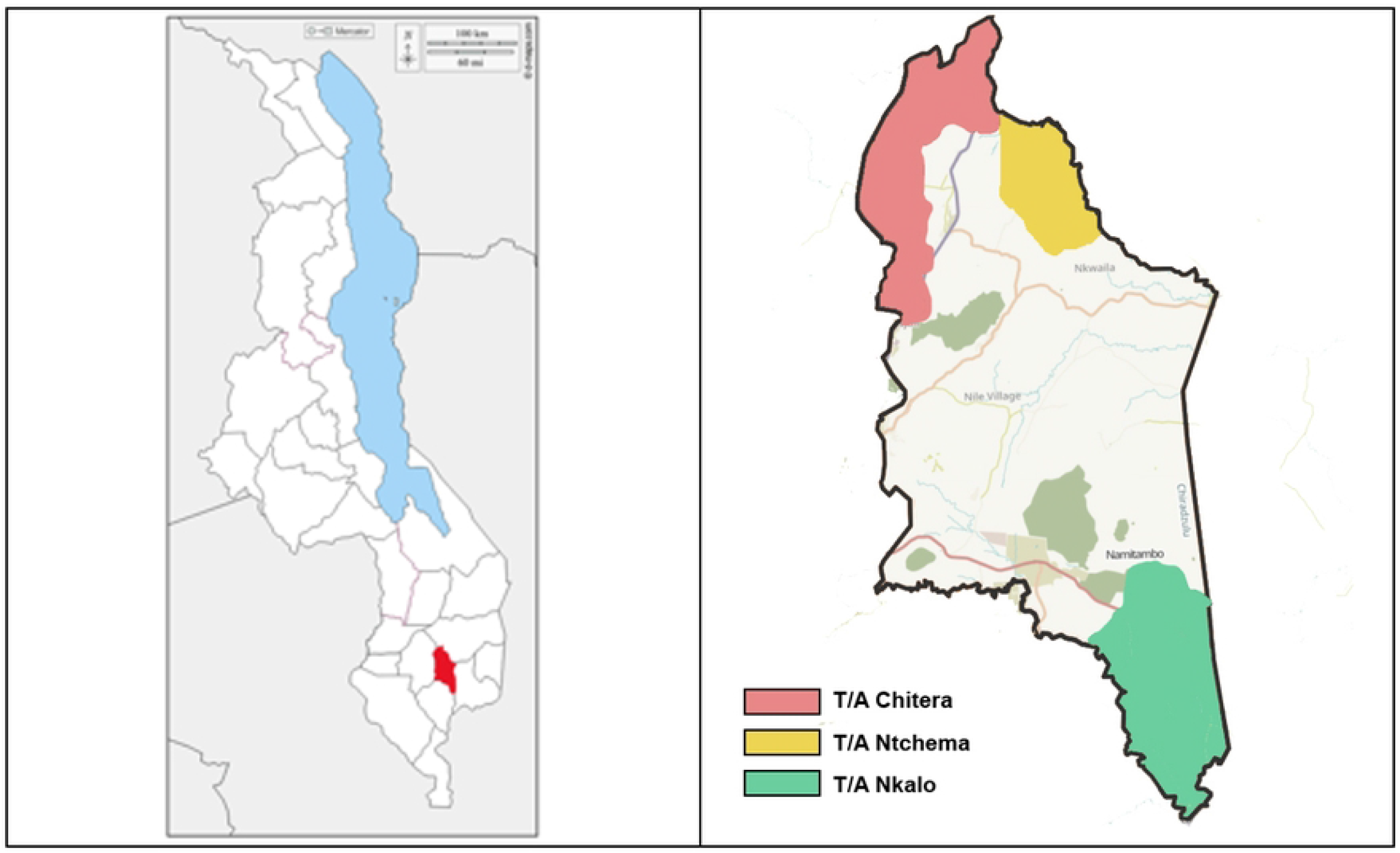
Map of Malawi focusing on Chiradzulu district (red), and (b) Map of Chiradzulu district depicting study areas: Red = Treatment 1: TA Chitera – CLTS + Care group arm, Yellow = Treatment 2: TA Ntchema – CLTS only arm and Green = Control arm: TA Nkalo

### Study Design

The study design is a controlled before-and-after (CBA) trial with two treatment arms (each with 20 villages/clusters) and a control group (30 villages) (Figure 2). CBA intervention designs are a non-randomized approach used to evaluate the impact of interventions (27,28). Advantages and disadvantages of the CBA study designs, also known as non-randomised cluster-controlled trials, have been discussed elsewhere (29).

**Figure 2.**
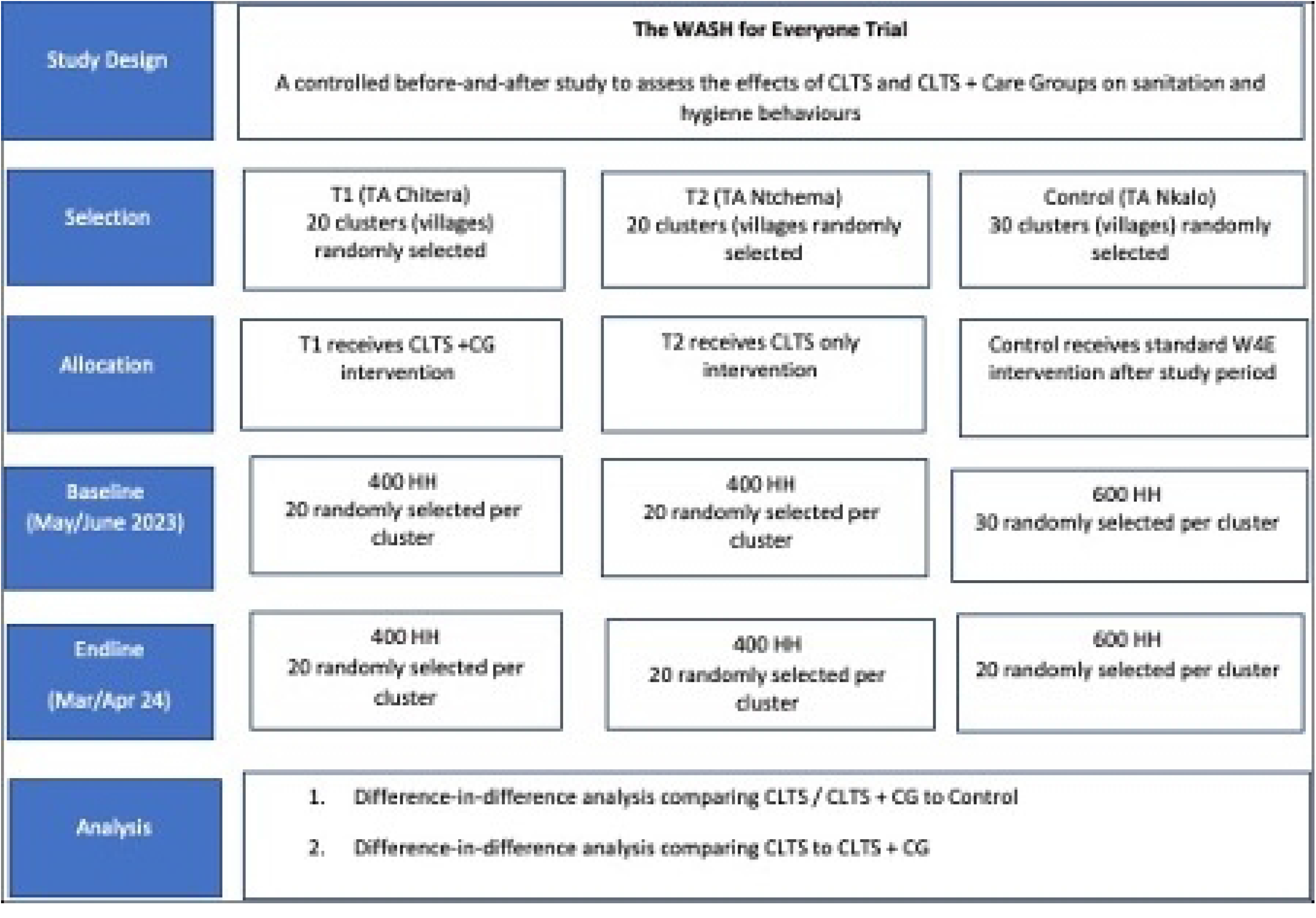
Study CONSORT diagram for a controlled before-and-after trial to assess the effectiveness of two interventions (CLTS only and CLTS and Care Group model) on rural sanitation coverage and use in Chiradzulu District, Malawi.

In our study, we selected three TAs (Figure 1) in Chiradzulu District, Malawi. TAs are 4^th^ level administrative units, with average population 34,000 in Chiradzulu (30). Details on the programme intervention are described below. For our trial, we selected two out of the five TAs scheduled to receive the full W4E intervention during the second year of programme implementation to align with our implementation. Each selected TA was randomly assigned to one of two intervention arms (CLTS or CLTS + care groups). We selected a third TA from the three TAs scheduled to receive the WASH for Everyone intervention in the third year of programme activities (Fig 2). This TA served as the control group for our study. While TAs are the unit of intervention assignment, villages (clusters) are the unit of analysis.

### Description of the intervention

The sanitation intervention included in the W4E programme uses CLTS, in line with the Malawi National Sanitation and Hygiene Strategy (31). As part of the W4E project, relevant district technical officers and community leaders (known natural leaders) implemented CLTS. Trained CLTS facilitators conducted the triggering sessions that include participatory activities, such as walk of shame, shit calculation and community sanitation mapping. The purpose of these activities is to trigger behavioural emotions, such as shame and disgust, so that community members understand the consequences of open defaecation. If successfully implemented, triggering sessions have the potential to stimulate community members to stop open defaecation and adopt improved sanitation practices, including construction and effective use of latrine facilities. For this trial, TA Ntchema (Treatment Arm 2) will receive the standard CLTS intervention (Table 1).

TA Chitera (Treatment Arm 1) will receive the standard CLTS intervention with the incorporation of Care Groups (CGs). CG leaders and cluster leaders support intervention delivery and local triggering sessions, specifically by facilitating CG meetings with CG households and conducting household follow-up visits. Care Groups are intended to extend the reach of CLTS behaviour change messaging, providing additional points of contacts with programme households. Care Groups also participate in post-triggering follow-up visits to households to assess sanitation coverage and use.

In line with CLTS values, the W4E project does not intend to provide any latrine construction or hygiene facility materials or financial subsidies to the households. It is the responsibility of the household owners to support themselves throughout the latrine and hygiene facility construction process.

### Selection of clusters and households

The primary sampling unit (clusters) for the study are clusters (also called villages) and represent the units of W4E delivery for CLTS. Communities from the three participating TAs were randomly selected from a list of communities obtained from the Chiradzulu District Health Office. Inclusion criteria for the selected communities include that the community is in one of the three selected Traditional Authorities that are part of the study area and have not yet received any exposure to the W4E project. Communities that were exposed to CLTS-related activities in the past 12 months and are outside the area of the three selected TAs are excluded from the study.

In early 2023, a list of all clusters in each study TA and their associated number of households was provided by the District Health Office. TA-specific median village size was calculated, and villages categorized as either above or below the TA-specific median. Villages in each TA-specific above and below median list were rank ordered according to a random number generated in Microsoft Excel and villages enrolled sequentially until the necessary number of villages were enrolled. If a cluster could not be located or if the village chief did not provide permission for data collection, the next cluster on the rank order list was enrolled.

Households are the secondary sampling unit and individuals living in households in the study area are the primary study population. Among selected communities (T1: n= 20; T2: n= 20; C: n= 30), systematic sampling procedures were followed to select up to 20 households per cluster. In selected households, we identified one adult resident (i.e. 18+ years old) to serve as the primary respondent, preferably the household head. Household samples are taken independently at baseline and endline. While study clusters will remain the same between baseline and endline, different households may be selected between baseline and endline.

### Data collection

Baseline, mid-line and end-line data collection

A baseline survey was conducted between April and May 2023, before W4E Year 2 implementation in Treatment TAs 1 and 2. An endline survey will be conducted before W4E Year 3 implementation in the control TA, with data collection scheduled for March -and April 2024. A structured questionnaire with closed-ended questions with pre-coded responses on mobile device on KOBO collect platform will be used to collect data on household membership, wealth index, sanitation and hygiene facilities, and child health. Further, the enumerators will conduct spot checks and record hygiene proxy measures, such as presence and state of a latrine, presence, location, and type of handwashing facility (including the availability of soap and water). The interviews will be conducted in Chichewa, the local language of Chiradzulu District. Chichewa-speaking enumerators with extensive training and expertise will administer the structured questionnaire.

To confirm availability of WASH infrastructure and hygiene practices, we will conduct structured observations in 350 randomly selected randomly selected households (i.e., 100 households from each treatment arm and 150 households from the control group) among the recruited 1,400 households. Specifically, the observations are intended to capture practices pertaining to presence of latrine and handwashing facility with soap, handwashing practice at critical times (i.e. before eating, before food preparation, after changing child nappy and after latrine use) and child faeces disposal. One observer will be placed at each selected household. As behaviours of interest mostly occur in the morning hours to noon (32), our observations will last four hours from 8:00am to 12:00 noon.

### Data management

Data collected using mobile devices will be uploaded directly online to KOBO daily. Only the PI, co-investigators, and study personnel with authorised access will have access to the online data. At the end of the data collection, the full dataset will be uploaded into STATA v18 (Stata Corp, College Station, TX) for analysis. Although no identifiable information will be recorded during the surveys, study personnel will review the final database and permanently delete any identifiable information inadvertently collected during surveys. All data is stored on encrypted, password-protected servers.

### Study outcomes and data analysis

Statistical analysis will be carried out at the individual or household levels with appropriate adjustment for clustering within villages (and within households for individual-level outcomes). Data will be analysed according to the TA / intervention assignment irrespective of whether the intervention was taken up fully, partly or not at all. Since this is an effectiveness study, only an intention-to-treat dataset will be maintained.

The primary estimates of the effectiveness of the intervention on primary and secondary outcomes will be based on a difference-in-difference analysis and models adjusted for design variables alone (TA, village population above or below TA-specific median); a fixed effect for village size (above or below the median) will be included in the model. All models will include a dummy variable for treatment arm (CLTS, CLTS+ or control) and data collection round (baseline and endline) as well as the interaction term of those two variables.

To account for clustering of observation, hierarchical mixed-effects models will be used for all analysis. For household-level outcomes or outcomes where there is only one respondent per household, mixed effects models will account for clustering by including a random effect at the village level. For outcomes with multiple respondents or data points per household (specifically sanitation use, and hand washing behaviour), an additional random effect will be added at the household-level.

All analysis of primary and secondary outcomes will be based on the difference-in-difference approach. The regression coefficient of the interaction term between treatment arm and data collection round will be used as the primary effect measure. For binary outcomes measure, we will use melogit with robust standard errors and regression coefficients exponentiated to estimate an Odds Ratio. For continuous outcomes, we will use meglm, with identity link and gaussian family.

### Analysis of primary outcomes

Primary outcome (presence of a sanitation facility) will be analysed using hierarchical mixed effects logistic regression with robust VCE. The results will be presented with 95% confidence intervals. Household specific presence of a sanitation facility is a binary household indicator based on observed latrine that the household reports as completed. Analyses will report on Odds Ratios (OR) and 95% confidence interval of each treatment arm compared to the control as well as the CLTS alone compared to CLTS+.

### Analysis of secondary outcomes

#### 1. Sanitation use

sanitation use is an individual-level binary indicator based on reported location last time each member of the household used a latrine.

#### 2. Safe disposal of child faeces

will be analysed as a mixed effect model (household, village, and cluster) of observed household disposal site of last defaecation for all children under the age of five years.

#### 3. Basic sanitation coverage

is a household-level indicator based on presence of an observed sanitation facility that meets JMP criteria for basic sanitation facilities, specifically an improved sanitation type.

#### 4. Sanitation-related Quality of Life (SanQoL-5)

SanQoL-5 index is an individual-level variable reported by only one participant per household. It is a continuous variable (0 – 1) based on the weighted score of responses to five questions on a 3-level frequency scale (33).

#### 5. Latrine quality

latrine quality is a composite index based on observed and reported characteristics of household latrines [8]. Specific indicators that will be used include binary indicators for the following:

##### a. Hygiene

i. excreta hygienically contained (observed)
ii. whether facility leaks or overflows waste at any time of the year (reported)

##### b. Accessibility

i. whether everyone in the household able to access and use the toilet at all times of the day and night (observed and reported)
ii. toilet facility easily accessible even to young children, elderly, and disabled people (reported)

##### c. Sustainability

i. whether sanitation facility has durable pit lining or is connected to septic tank (observed and reported)
ii. whether pit latrine or septic tank has ever been emptied (reported)

##### d. Use

i. pit not full (observed)
ii. child faeces go into latrine or no children living on plot (observed and reported) Items will be combined using principal components analysis (PCA) and iteratively refined using standard approaches to index development. Outcomes will be analyzed as a continuous variable based on PCA score.

#### 6. Basic handwashing facility

presence of a handwashing where both soap and water are available is a household-level binary variable. This will be analyzed in two ways: a) both observed and reported HWF with both soap and water available at the time of data collection and b) observed HWF with soap and water.

#### 7. Handwashing behaviour

A binary outcome measure based on structured observation data. Structured observation data will be used to identify all pre-defined hand hygiene opportunities (e.g., before food preparation, before eating, before feeding child, after using a latrine, after cleaning a child, and after being in contact with an animal) and associated hand hygiene (0 = no hand hygiene or hand hygiene with water only; 1 = hands washed with soap). Analysis will be conducted at the event-level with adjustment for repeated observations within the same household.

### Adjustment for covariates

Results will present two sets of outcome measure. First, we will report on all outcome measures adjusted for design variables (village above or below TA-specific median) and models also adjusted for a priori defined covariates and design variables. We will explore differences between the design-adjusted and covariate-adjusted models but will consider the covariate adjusted models as the primary effect estimates.

Covariates have been selected based on hypothesized relationships that could confound the relationship between intervention exposure and primary and secondary outcome measures.

At the respondent level, gender and primary education (less than vs completed primary) will be included. While the following covariates will be included at household-level: household size, household economic status based on PCA of household assets, any member of the household experiencing a disability as defined by the Washington Group (any functional disability for a member of the household greater than two on the functional disability assessment) and household reports a water source that is located on site or on plot (34). At the community-level, we will look at village having an improved road as reported by Village Chief at time of baseline enrolment. Improved road is used as a proxy measure of village accessibility to both the intervention and markets.

### Missing data

It is likely that some missing outcome data will be encountered, especially of individual-level self-reported outcomes. The patterns of missingness of variables will be tabulated to describe and compare the extent of missingness of any affected variable between study arms. No adjustment will be made for missing data.

### Outliers

Unusual values and potential outliers will be flagged and queried. Unlikely values will be dropped and treated as missing data in the main analysis. A sensitivity analysis will be conducted which includes potential outliers (but not unlikely values).

### Multiple comparisons

The number of primary outcomes that will be tested for significant differences between arms is small; thus, no formal adjustment for multiple comparisons will be made.

#### Ethics

The study protocol, which includes data collection tools, participant information sheets and consent forms, have been approved by the National Commission for Science and Technology (P01/23/718) in Malawi. Further, consent was obtained from the Chiradzulu District Council and community leaders. Informed written consent was obtained from all study participants recruited into the study prior to data collection. The study was registered with clinicaltrials.gov (NCT05808218).

## Results

### Baseline data

Table 2 presents the demographic details of the 1,400 survey respondents across the three TAs. Four hundred respondents were sampled from Treatment TA 1, 400 from Treatment TA 2, and 600 from the control TA. Baseline characteristics were distributed evenly between the trial arms. Across the three TAs, most survey respondents were female (83%), had at least some primary education (67%), and were married (68%). The median household size was four members. Approximately half (49%) of households had a child under the age of five. Households were equally distributed across wealth quintiles, with approximately 20% of households in each of the five quintiles across the three TAs. Six percent of households had at least one member living with a disability.

**Table 2.**
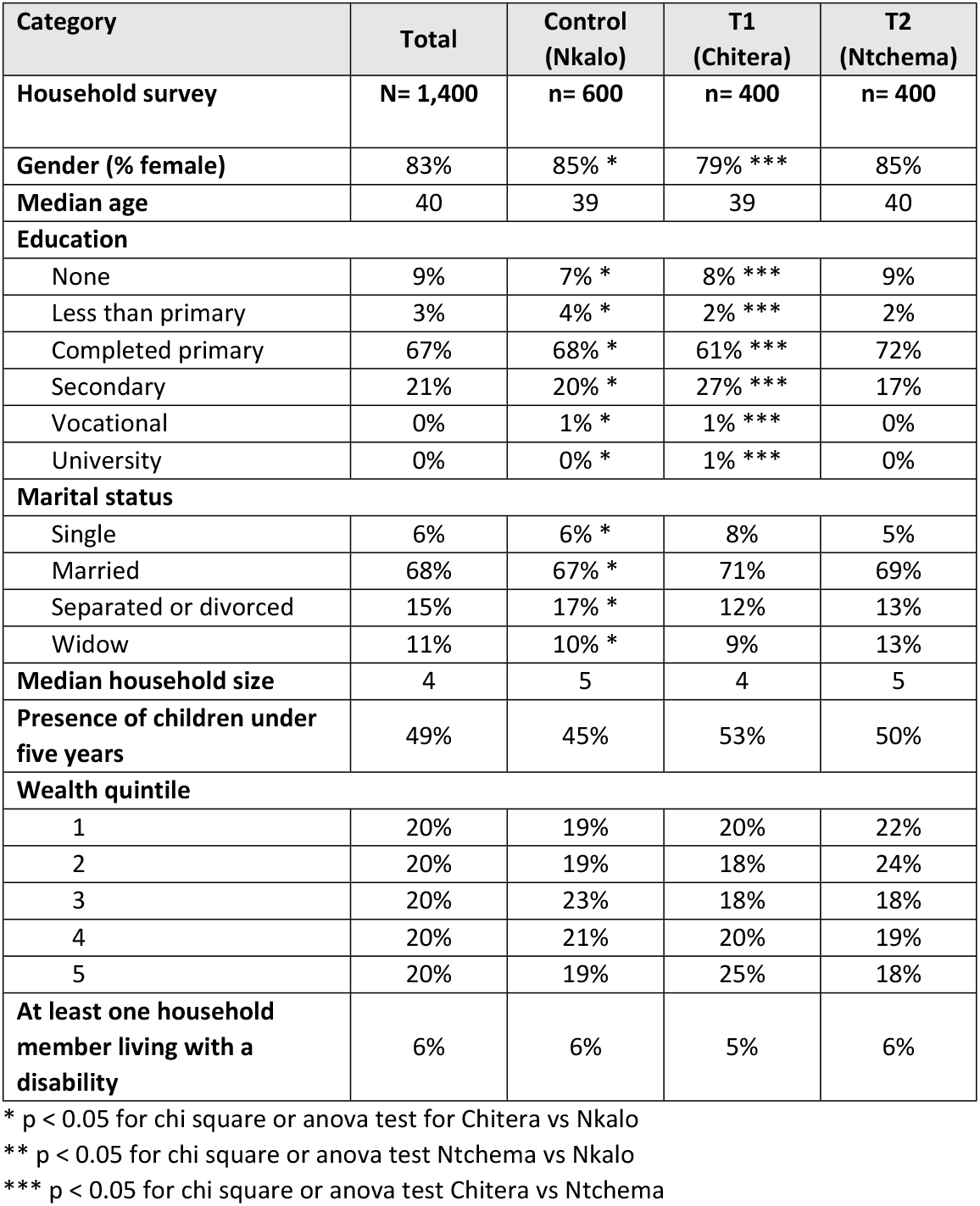
Description of baseline respondent characteristics.

At baseline, most households (78%) had an unimproved sanitation facility as defined by JMP (e.g., a pit latrine without a slab). Specifically, most households (76%) had a pit or twin pit latrine without slab and in yard or plot (68%). The median number of households amongst those sharing one latrine was 3. Ninety four percent of households reported all household members using a sanitation facility for the last defaecation event. Among the households with a sanitation facility, almost all (98%) reported that they regularly use their sanitation facility. The mean sanitation quality of life (SanQoL) is 0.62. Most households used an improved water source as defined by <u>JMP</u> (98%) (Table 3). More specifically, most households (94%) used a borehole as their main water source. Twenty-five minutes was the median round-trip time to collect water.

**Table 3.** Description of outcomes at baseline.

| Category | Total | Control (Nkalo) | T1 (Chitera) | T2 (Ntchemba) |
| --- | --- | --- | --- | --- |
| Household survey | **N= 1,400** | **n= 600** | **n= 400** | **n= 400** |
| Improved water source (JMP) | 98% | 98% | 97% | 98% |
| At least basic drinking water (JMP) | 70% | 78% * ** | 63% | 65% |
| Report treated water at least once a day | 63% | 57% * ** | 64% | 70% |
| Sanitation (JMP) |  |  |  |  |
| At least basic | 7% | 4% * ** | 11% *** | 7% |
| Limited | 11% | 9% * ** | 16% *** | 8% |
| Unimproved | 78% | 81% * ** | 70% *** | 83% |
| No facility | 4% | 6% * ** | 4% *** | 2% |
| Number of households sharing one latrine (amongst those sharing), Median (IQR) | 2 (2, 3) | 2 (2, 3) | 3 (2, 3) | 2 (2, 3) |
| All members of the household (aged ≥5) reported using latrine for last defecation | 96% | 94% * ** | 97% | 98% |
| SanQoL-5 index, mean (SD) | .62 (0.25) | .59 (0.26) | .64 (0.26) | .64 (0.24) |
| Reported child faeces disposal (n= 715) |  |  |  |  |
| Safe disposal |  | 90% | 88% | 90% |
| Unsafe disposal |  | 10% | 12% | 10% |
| Handwashing station available (reported or observed) |  |  |  |  |
| Basic (handwashing facility with soap and water) | 7% | 10% * | 4% *** | 8% |
| Limited | 49% | 52% | 42% | 52% |
| No handwashing facility | 44% | 39% * | 54% *** | 41% |

\* p < 0.05 for chi square or anova test for T1 vs Control
\*\* p < 0.05 for chi square or anova test T2 vs Control
\*\*\* p < 0.05 for chi square or anova test T1 vs T2

Over half of respondents (56%) had access to a handwashing station, either observed or reported (Table 3). However, only 7% of households had a handwashing facility with reported soap and water available within the household and 6% with observed soap and water available at the handwashing facility.

Three hundred and thirty hand hygiene structured observations were conducted in Treatment TA 1, 295 in Treatment TA 2, and 474 in the control TA (Table 4). The median number of observed hand hygiene opportunities per household was three. Most households did not practice hand hygiene or wash hands with water at specific hand hygiene junctures (e.g., after using the toilet, before food preparation, before eating, before feeding child, and after being in contact with an animal) (Table 4). A total of 73 child faeces disposal structured observations were conducted across the three TAs, with 26 in T1, 18 in T2, and 29 in the control. Approximately half of households safely disposed of child faeces (i.e., put or rinsed the faeces in the latrine) in the three TAs (Table 4).

**Table 4.** Hand hygiene and child faeces disposal structured observations.

| Category | Total | Nkalo | Chitera | Ntchema |
| --- | --- | --- | --- | --- |
| **Hand hygiene structured opportunities** | **N= 1,099** | **n= 474** | **n= 330** | **n= 295** |
| Proportion handwashing with soap and water | 3% | 3% | 5% *** | 2% |
| Proportion handwashing with water only | 46% | 49% | 39% *** | 50% |
| Proportion handwashing with other materials (i.e. ash) | 0% | 0% | 0% *** | 0% |
| Proportion no handwashing | 51% | 50% | 56% *** | 48% |
| **Child faeces disposal structured observations** | **N= 73** | **n= 29** | **n= 26** | **n= 18** |
| Safe disposal | 49% | 45% | 54% | 50% |
| Unsafe disposal | 51% | 55% | 46% | 50% |

\* p < 0.05 for chi square or anova test for T1 vs Control
\*\* p < 0.05 for chi square or anova test T2 vs Control
\*\*\* p < 0.05 for chi square or anova test T1 vs T2

## Discussion

This paper presents the study protocol and baseline findings of a community-based intervention promoting the integration of the CG model into the CLTS approach, given that CLTS alone is reportedly not effective enough to ensure sustained uptake of sanitation and hygiene behaviours. Studies have shown the effectiveness of using the CG model in promoting community health interventions, mainly nutrition programmes (20,35–38). However, few have been conducted which integrate CLTS and the CG model. To address this gap, we developed an innovative intervention approach that integrates the CG model into the standard CLTS approach. The CG model is community-level government-recognised approach that can potentially support the already existing informal - natural leaders in the delivery of CLTS, both pre- and post-ODF status attainment. This would strengthen CLTS sustainability since care group members are trained to continuously monitor a specific group of households post-ODF attainment, while natural leaders would only follow households up to community’s attainment of ODF status (9,13,39,40).

### Baseline survey results

Our baseline results indicate that the study is conducted in a rural area where literacy level is moderate and the quality of housing relatively poor. Some households had no latrines, while traditional unimproved latrines (e.g., pit latrines without slab), which are prone to collapsing during rainy season, were common. Relatively high sanitation coverage in the communities indicates the short-term success of CLTS, however, other TAs in Chiradzulu district reverted to OD after attainment of ODF status. Thus, the district provided a suitable environment for this study to assess alternative approaches for promoting sustainable sanitation interventions.

Handwashing with soap is low in low- and middle-income countries, including Malawi (41–45). Similar findings were observed in our baseline results. Access to easy to use and effective handwashing facilities was limited among the study population as most households did not have a dedicated handwashing facility. Observations at critical times indicated a challenge with soap availability for hand washing. As such, nearly half of the study participants did not use soap during handwashing.

### Comparison of baseline characteristics among trial arms

The baseline results indicate some imbalance between trial arms on self-reported water treatment practices. Treatment of household water was more frequently reported among the control group compared to the treatment groups. A possible explanation for the imbalance could be that immediately before baseline data collection, other non-governmental organisations promoted the use of safe water through the drilling of boreholes and installation of chlorine dispensers in all the water points in the control communities. This included the installation of chlorine dispensers in both treatment groups, although a similar trend was not observed in responses. In terms of sanitation, we observed that the control arm had the highest coverage of latrines without a slab, while more latrines with a slab were observed in T2. We could not find an explanation for this difference and expect it to be by chance.

This study has several limitations. Structured observations were used to collect the data. This approach has limitations as the presence of the observer can influence the behaviour of the person under observation (46). To minimise bias, study participants were not informed about the observed hygiene behaviours, though they were generally informed about the purpose of the study. Nonetheless, structured observations remain the gold standard for measuring hygiene behaviours (47,48). Treatment arms and the control were purposively selected based on the presence of care groups and CLTS programme for the treatment arms rather than random assignment. The control arm was selected based on the absence of care groups and CLTS. Systematic random sampling was used to select study households in each arm.

## Conclusion

We highlight the study design and baseline results from an ongoing BCA trial implemented in rural households in Chiradzulu District, Malawi. The study, implemented within a district-wide WASH programme, assesses the effectiveness of CLTS alone against CLTS combined with the CG model for improving sanitation coverage and sustained uptake. The baseline observations indicate a balanced distribution of potential demographic confounders in the trial arms with a slight variation on some WASH proxy measures. We expect to publish the trial findings in early 2025.

## Data Availability

All data referred to within this study will be available in a repository upon acceptance.

## Author’s contributions

The trial was designed by RD, KC and TM. Sample size estimates and randomization procedures were guided by MV. The trial is coordinated by KC with input from RD, TM, IR, MP and CM. Baseline survey activities were coordinated by MP and BW. Analysis of baseline survey data was performed by CM. KC and RD drafted the manuscript. All co-authors contributed critically to the manuscript and approved the final version.

## Supplementary materials

**Table A1.** Description of baseline respondent characteristics.

| Variable | Total<br>N= 1,400 |  | Nkalo (control)<br>N = 600 |  | Chitera (T1)<br>N= 400 |  | Ntchema (T2)<br>N= 400 |  |
| --- | --- | --- | --- | --- | --- | --- | --- | --- |
|  | n | % | n | (%) | n | % | n | % |
| **Gender** | 1,167 | 83% | 510 | 85% * | 317 | 79% *** | 340 | 85% |
| **Median respondent age** | 1,400 | 40 | 600 | 39 | 400 | 39 | 400 | 40 |
| **Education** |  |  |  |  |  |  |  |  |
| None | 107 | 9% | 44 | 7% * | 30 | 8% *** | 33 | 9% |
| Less than primary | 43 | 3% | 25 | 4% * | 9 | 2% *** | 9 | 2% |
| Completed primary | 940 | 67% | 405 | 68% * | 246 | 61% *** | 289 | 72% |
| Secondary | 296 | 21% | 119 | 20% * | 110 | 27% *** | 67 | 17% |
| Vocational | 6 | 0% | 2 | 1% * | 3 | 1% *** | 1 | 0% |
| University | 8 | 1% | 5 | 0% * | 2 | 1% *** | 1 | 0% |
| **Marital status** |  |  |  |  |  |  |  |  |
| Single | 85 | 6% | 33 | 6% * | 33 | 8% | 19 | 5% |
| Married | 958 | 68% | 37 | 67% * | 20 | 71% | 15 | 69% |
| Separated or divorced | 208 | 15% | 103 | 17% * | 48 | 12% | 57 | 13% |
| Widowed | 149 | 11% | 62 | 10% * | 37 | 9% | 50 | 13% |
| **Median household size** | 1,400 | 4 | 600 | 5 | 400 | 4 | 400 | 5 |
| **Presence of children under five years** | 171 | 49% | 68 / 150 | 45% | 53 / 100 | 53% | 50 / 100 | 50% |
| **Wealth quintile** |  |  |  |  |  |  |  |  |
| 1 | 281 | 20% | 115 | 19% | 80 | 20% | 86 | 22% |
| 2 | 279 | 20% | 114 | 19% | 71 | 18% | 94 | 24% |
| 3 | 280 | 20% | 135 | 23% | 72 | 18% | 73 | 18% |
| 4 | 280 | 20% | 123 | 21% | 81 | 20% | 76 | 19% |
| 5 | 280 | 20% | 113 | 19% | 96 | 24% | 71 | 18% |
| **At least one household member living with a disability** | 72 | 5% | 31 | 6% | 18 | 5% | 23 | 6% |

**Table A2.** Description of outcomes at baseline.

| **Variable** | **Total** |  | **Nkalo (control)** |  | **Chitera (T1)** |  | **Ntchema (T2)** |  |
| --- | --- | --- | --- | --- | --- | --- | --- | --- |
|  | **n / N** | **%** | **n / N** | **%** | **n / N** | **%** | **n / N** | **%** |
| **Improved water source (JMP)** | 1,364 / 1,395 | 98% | 587 / 598 | 98% | 384 / 397 | 97% | 393 | 98% |
| **At least basic drinking water (JMP)** | 980 / 1,395 | 70% | 466 / 598 | 78% * ** | 253 / 397 | 63% | 261 / 400 | 65% |
| **Report treated water at least once a day** | 876 / 1,400 | 63% | 342 / 600 | 57% * ** | 255 / 400 | 64% | 279 / 400 | 70% |
| **Sanitation (JMP)** |  |  |  |  |  |  |  |  |
| At least basic | 97 / 1,400 | 7% | 26 / 600 | 4% * ** | 43 / 400 | 11% *** | 28 / 400 | 7% |
| Limited | 149 / 1,400 | 11% | 55 / 600 | 9% * ** | 64 / 400 | 16% *** | 30 / 400 | 8% |
| Unimproved | 1,094 / 1,400 | 78% | 483 / 600 | 81% * ** | 278 / 400 | 70% *** | 333 / 400 | 83% |
| No facility | 60 / 1,400 | 4% | 36 / 600 | 6% * ** | 15 / 400 | 4% *** | 9 / 400 | 2% |
| **Number of households sharing one latrine (amongst those sharing), Median (IQR)** | 825 | 2 (2, 3) | 396 | 2 (2, 3) | 203 | 3 (2, 3) | 226 | 2 (2, 3) |
| **All members of the household (aged ≥5) reported using latrine for last defecation** | 5,183 / 5,386 | 96% | 2,201 / 2,338 | 94% | 1,480 / 1,520 | 97% *** | 1,502 / 1,528 | 98% |
| **SanQoL-5 index, mean (SD)** | 1,400 | 0.62 (0.25) | 600 | 0.59 (0.26) | 400 | 0.64 (0.26) | 400 | .64 (0.24) |
| **Reported child faeces disposal** |  |  |  |  |  |  |  |  |
| Safe disposal | 641 / 715 | 90% | 263 / 291 | 90% | 184 / 209 | 88% | 194 / 215 | 90% |
| Unsafe disposal | 74 / 715 | 10% | 28 / 291 | 10% | 25 / 209 | 12% | 21 / 215 | 10% |
| **Handwashing station available (reported or observed)** |  |  |  |  |  |  |  |  |
| Basic (HWF with soap and water) | 104 | 7% | 59 / 600 | 10% * | 15 / 400 | 4% *** | 30 / 400 | 8% |
| Limited | 683 | 49% | 309 / 600 | 52% | 168 / 400 | 42% | 206 / 400 | 52% |
| No handwashing facility | 613 | 44% | 232 / 600 | 39% * | 217 / 400 | 54% *** | 164 / 400 | 41% |

**Table A3.**
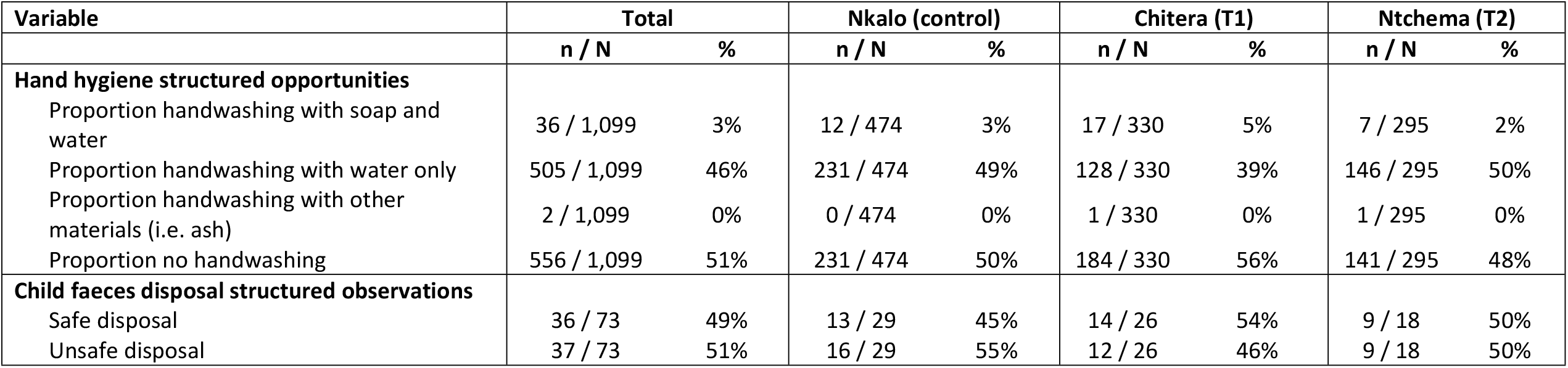
Hand hygiene and child faeces disposal structured observations.

| Variable | Total |  | Nkalo (control) |  | Chitera (T1) |  | Ntchema (T2) |  |
| --- | --- | --- | --- | --- | --- | --- | --- | --- |
|  | n / N | % | n / N | % | n / N | % | n / N | % |
| **Hand hygiene structured opportunities** |  |  |  |  |  |  |  |  |
| Proportion handwashing with soap and water | 36 / 1,099 | 3% | 12 / 474 | 3% | 17 / 330 | 5% | 7 / 295 | 2% |
| Proportion handwashing with water only | 505 / 1,099 | 46% | 231 / 474 | 49% | 128 / 330 | 39% | 146 / 295 | 50% |
| Proportion handwashing with other materials (i.e. ash) | 2 / 1,099 | 0% | 0 / 474 | 0% | 1 / 330 | 0% | 1 / 295 | 0% |
| Proportion no handwashing | 556 / 1,099 | 51% | 231 / 474 | 50% | 184 / 330 | 56% | 141 / 295 | 48% |
| **Child faeces disposal structured observations** |  |  |  |  |  |  |  |  |
| Safe disposal | 36 / 73 | 49% | 13 / 29 | 45% | 14 / 26 | 54% | 9 / 18 | 50% |
| Unsafe disposal | 37 / 73 | 51% | 16 / 29 | 55% | 12 / 26 | 46% | 9 / 18 | 50% |

